# Comparative genomic analysis reveals shared and distinct mechanisms of nasal polyps and chronic rhinosinusitis

**DOI:** 10.64898/2026.04.07.26350325

**Authors:** Shuai Yuan, John C McVey, Katherine Hartmann, Sarah Abramowitz, Jakob Woerner, Gabrielle Shakt, Renae Judy, Jennifer E. Douglas, Benjamin F. Voight, Michael A. Kohanski, Noam A. Cohen, Michael G. Levin, Scott M. Damrauer

## Abstract

**Background:** Chronic rhinosinusitis (CRS) and nasal polyps (NP) are closely related inflammatory airway diseases, and their co-occurrence is often associated with more persistent symptoms, frequent recurrence, and substantial respiratory morbidity. However, the extent to which CRS without and with NP (CRSsNP and CRSwNP) share genetic susceptibility—and which genetic mechanisms are disease-specific—remains poorly characterized.

**Methods:** We conducted cross-population genome-wide association meta-analyses of overall CRS (including both CRSwNP and CRSsNP) and NP (a proxy for CRSwNP) using data from six biobanks. We estimated genome-wide genetic correlations between overall CRS, CRSwNP, and a spectrum of respiratory diseases. We applied five complementary gene-prioritization strategies to nominate CRS- and CRSwNP-associated genes and performed pathway enrichment analyses to infer implicated biological processes. For CRSwNP, we integrated single-cell transcriptomic data to characterize cell-type–specific expression of prioritized genes and used stratified LD score regression to quantify heritability enrichment across immune and epithelial annotations. To delineate shared versus disease-specific genetic signals, we performed three comparative analyses—local genetic correlation, CRSwNP-CRS colocalization, and genomic structural equation modeling. Finally, we performed proteome-wide Mendelian randomization to identify circulating proteins with putative causal effects on CRS and CRSwNP.

**Results:** This GWAS meta-analysis identified 96 genome-wide significant loci for CRSwNP and 41 for overall CRS, prioritizing 92 and 39 candidate genes, respectively. CRSwNP and overall CRS showed shared genetic susceptibility (*r_g_* = 0.59; *P* = 6.8 × 10⁻¹□), while CRS exhibited broader genetic correlations across multiple respiratory disorders. Pathway analyses consistently implicated immune signaling albeit with disease-specific emphases and lipid-metabolism networks. Single-cell analyses demonstrated distinct expression of CRSwNP-prioritized genes across nasal epithelial and immune cell clusters, and immune annotations explained more CRSwNP heritability (enrichment score = 4.1; *P* = 0.010) than epithelial annotations (2.5; *P* = 0.072). Comparative genetic analyses highlighted multiple shared loci—including *BACH2, CD247, FADS2, FOXP1, FUT2, GPX4, IL7R, NDFIP1, RAB5B, RORA, SMAD3, TSLP* —as well as 3 CRSwNP-specific and 6 CRS-specific loci. Proteome-wide MR identified 10 and 8 putatively causal circulating proteins for CRSwNP and overall CRS, respectively, with protein TNFSF11, IL2RB, and STX4 associated with both conditions.

**Conclusions:** This multi-population GWAS meta-analysis expanded genetic discovery for CRS and CRSwNP and showed substantial shared liability with distinct disease-specific components. Immune components explained a larger proportion of CRSwNP heritability than epithelial annotations, reinforcing the primacy of immune-driven mechanisms in polyp disease.

## Introduction

Chronic rhinosinusitis (CRS) is a common inflammatory disorder of the nose and paranasal sinuses defined by persistent sinonasal symptoms ≥12 weeks in duration plus objective evidence of inflammation on endoscopy or imaging. It affects approximately 5–12% of the general population and imposes substantial quality-of-life and healthcare burdens (1). A major clinical distinction within CRS is the presence (CRSwNP) or absence (CRSsNP) of nasal polyps, with polyp disease affecting around 1-4% of the population (2)). Nasal polyps are benign inflammatory growths of sinonasal mucosa that are strongly linked to type 2 inflammation and comorbid asthma, and CRSwNP is characterized by treatment-recalcitrant symptoms and a propensity for recurrence (3,4).

Genome-wide association studies (GWAS) have begun to map inherited susceptibility to CRS and NP (or CRS with NP [CRSwNP]). Early studies were underpowered, but implicated involvement of immune-related loci, including signals in the *HLA* region for CRSwNP (5). Larger-scale work then identified more definitive associations: a landmark GWAS in Icelandic and UK datasets reported a protective loss-of-function variant in *ALOX15* for both NP and CRS, suggesting lipid mediator biology as a causal pathway rather than a downstream correlate of inflammation (6). In addition, a two-stage GWAS incorporating imaging-derived CRS sub-phenotypes found that genetic associations differed between imaging-defined patterns (e.g., localized versus diffuse opacification), consistent with etiologic heterogeneity within clinically defined CRS (7). Most recently, biobank-scale meta-analyses integrating FinnGen and UK Biobank expanded locus discovery and reported substantially more loci for CRSwNP than for CRS without NP (CRSsNP), reinforcing differences in genetic architecture across CRS subtypes (8).

Despite this progress, important knowledge gaps remain. Prior GWASs have often been underpowered for overall CRS relative to NP (or CRSwNP) and have relied heavily on single-ancestry discovery. Critically, most studies have analyzed CRS and NP (or CRSwNP) in parallel rather than comparatively, limiting the ability to separate loci reflecting shared liability to chronic sinonasal inflammation from loci that are specific to polyp formation, tissue remodeling, or severe endotypes. To address these gaps, we performed a multi-ancestry GWAS meta-analysis of NP (a proxy for CRSwNP) and overall CRS (a phenotype that captures both CRSwNP and CRSsNP) and directly interrogated both shared and disease-specific genetic architecture.

## Results

The multi-population GWAS meta-analysis comprised 17,046 individuals with CRSwNP and 1,573,214 without, together with 97,519 individuals with overall CRS and 1,612,702 without, sampled across six population groups (**Table S1**). Although both traits exhibited moderate genomic inflation (λ_GC_ = 1.21 for CRSwNP; λ_GC_ = 1.34 for CRS), LDSC intercepts of 1.10 (SE = 0.01) and 1.08 (SE = 0.02), with attenuation ratios of 25.5% and 21.0%, respectively, indicate that most of this inflation reflects true polygenic architecture rather than residual bias. On the liability scale, SNP heritability was estimated at 17.34% (SE = 2.21%) for CRSwNP and 4.45% (SE = 0.29%) for CRS. CRSwNP and CRS exhibited a moderate-to-strong genome-wide genetic correlation (*r_g_* = 0.59; *P* = 6.8 × 10⁻¹□).

### Multi-ancestry GWAS meta-analysis uncovered 96 risk loci for CRSwNP and 41 for overall CRS

We detected 96 genome-wide significant loci for CRSwNP and 41 for overall CRS (**Table S2**). In CRSwNP, lead variants at *ALOX15*, *KDM4C*, *NELFE*, *RIC1*, *IL33*, and *HLA-DQA1* exhibited per-allele odds ratios exceeding 1.3 (**Figure 1a; Table S2**), whereas for CRS the strongest effects were observed at *HLA-C, SLC39A7*, and *ZSCAN16* (**Figure 1b; Table S3**). Across all loci, we prioritized 92 genes for CRSwNP and 39 for CRS. Among the CRSwNP candidates, 13 genes were supported by three independent prioritization methods, and 13 genes harbored variants within credible sets predicted to be loss-of-function or missense variants (functional annotation) (**Figure 1c; Table S4**). For CRS, 5 genes met the three-method criterion and 8 were flagged by functional annotation (**Figure 1d; Table S5**).

**Figure 1.**
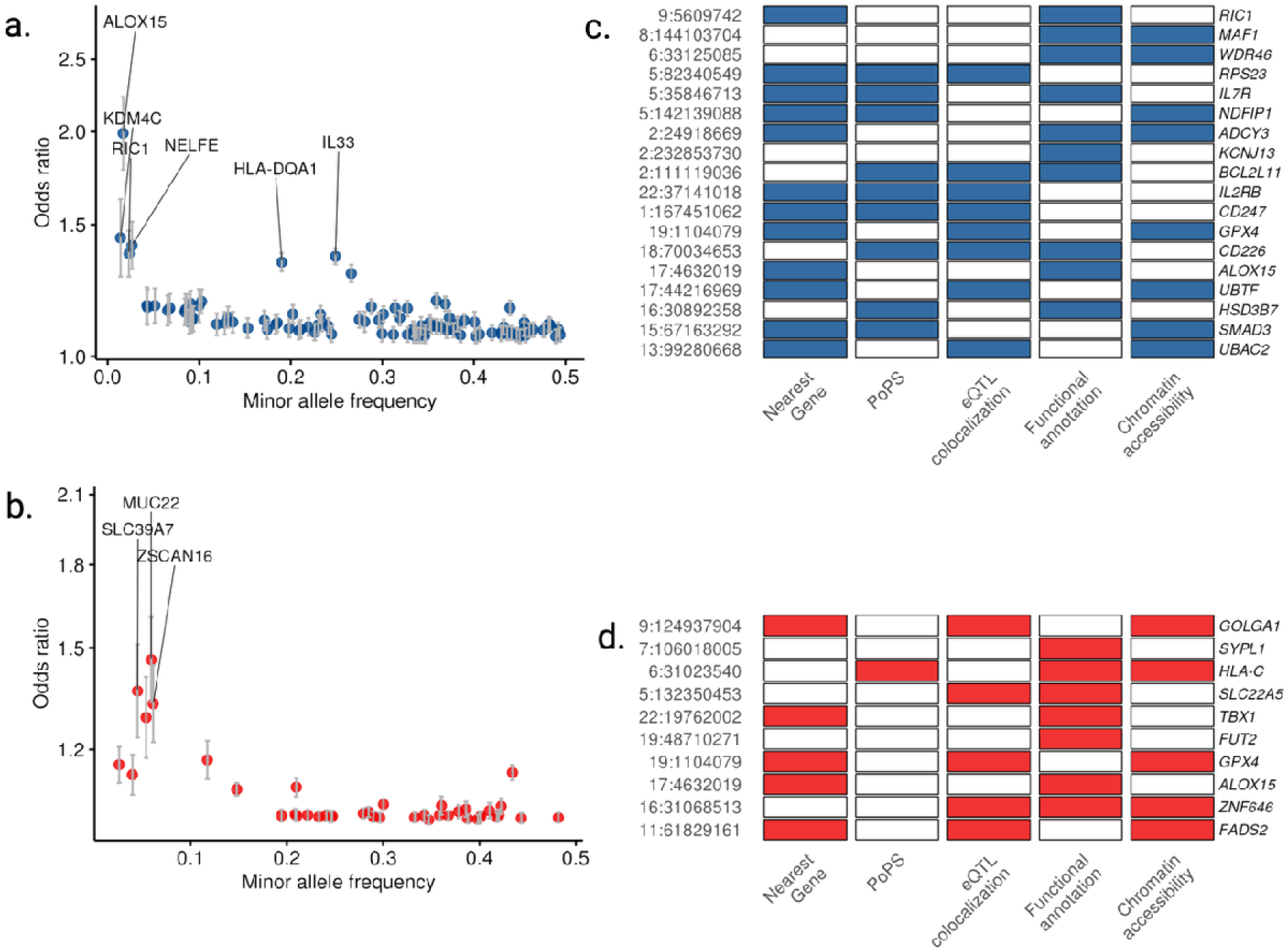
Identification and prioritization of genetic loci for chronic rhinosinusitis with nasal polyps (CRSwNP) and overall chronic rhinosinusitis (CRS). (A) Minor allele frequency (MAF) versus odds ratio (OR) for lead single-nucleotide polymorphisms (SNPs) associated with CRSwNP. (B) MAF versus OR for lead SNPs associated with overall CRS. Variants with OR > 1.3 are labeled. (C) Prioritized genes at CRSwNP loci based on multiple lines of evidence: PoPS (polygenic priority score), eQTL colocalization, pQTL overlap, chromatin interaction (Hi-C), and functional annotation. (D) Prioritized genes at overall CRS loci using the same methods. Only genes supported by ≥ 3 prioritization approaches or direct functional annotation are shown.

### CRSwNP and overall CRS share genetic architecture with multiple respiratory disorders

In a wide-angle genetic correlation experiment with 70 respiratory diseases in MVP, CRSwNP exhibited significant correlations with 16 endpoints after Bonferroni correction (*P*_<D0.05; **Table S6**), most prominently with asthma with exacerbation (*r*_₉_ = 0.57, *P* = 1.1 × 10⁻□). In contrast, CRS showed significant correlations with 48 respiratory outcomes (*P* < 0.05; **Table S7**), including bronchitis (*r*_₉_ = 0.67, P = 1.0 × 10⁻²□), asthma (*r*₉ = 0.55, P = 9.3 × 10⁻²³), and chronic airway obstruction (*r*_₉_ = 0.40, P= 7.0 × 10⁻³²).

### Pathway enrichment underscores immune signaling and lipid metabolism in CRSwNP and overall CRS

Using the 92 CRSwNP-prioritized genes, Reactome pathway analysis identified 17 significantly enriched pathways (**Figure 2a; Table S8**), and Gene Ontology (GO) biological process enrichment revealed 92 significant terms (**Figure 2b; Table S9**). CRSwNP genes were particularly overrepresented in cytokine-mediated signaling—most notably interleukin-4 and interleukin-13 pathways—and in RUNX3-related transcriptional programs. In parallel, analysis of the 39 CRS-prioritized genes uncovered 18 enriched Reactome pathways (**Figure 2c; Table S10**) and 48 significant GO biological processes (**Figure 2d; Table S11**), with dominant signatures in T-cell–mediated immune responses and interleukin-1 signaling. Notably, lipid metabolism pathways—especially those involving eicosapentaenoic and arachidonic acid—were enriched in both CRSwNP and CRS gene sets, suggesting shared metabolic contributions to disease pathogenesis.

**Figure 2.**
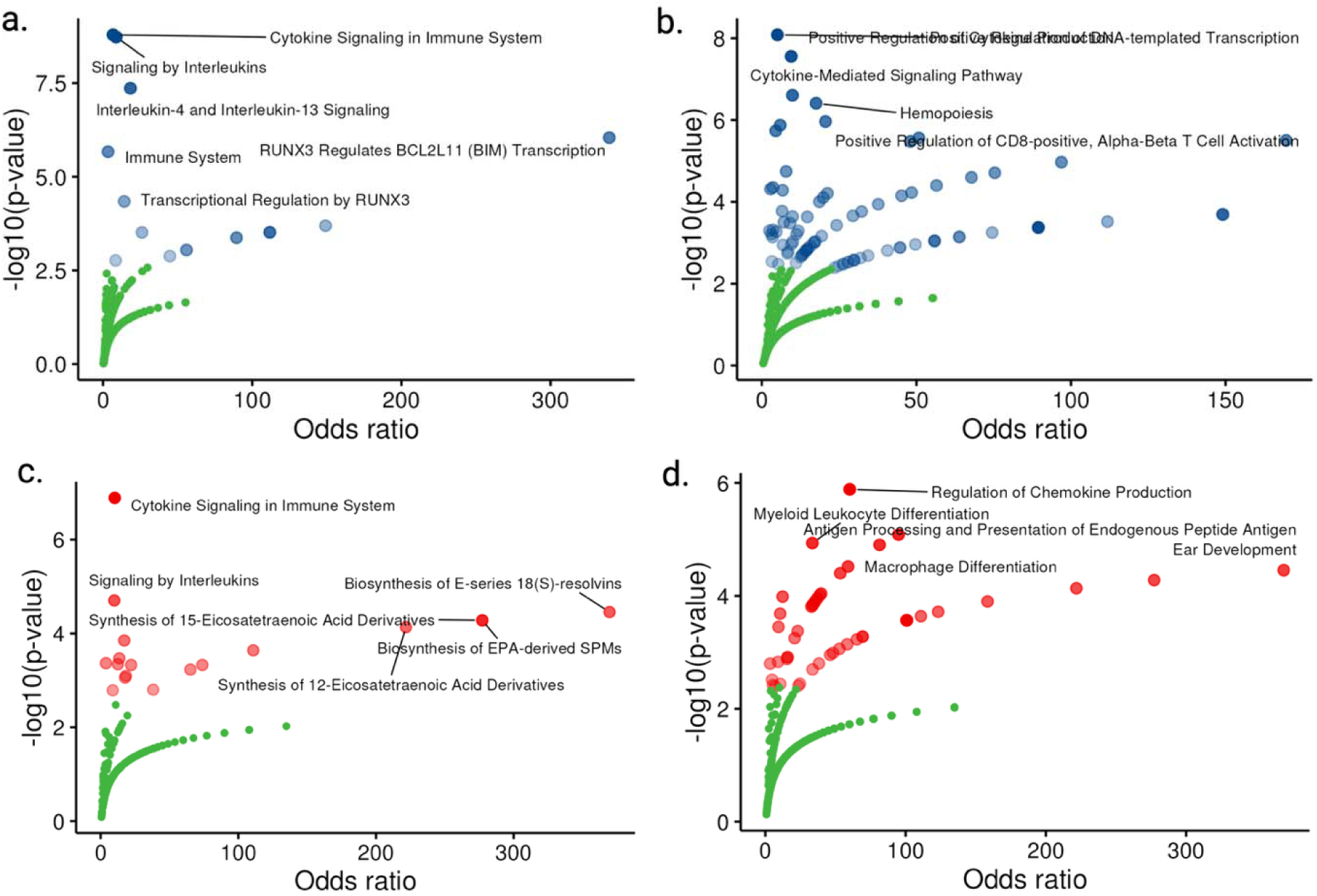
Enrichment of Reactome and Gene Ontology pathways among genes prioritized for chronic rhinosinusitis with nasal polyps (CRSwNP) and chronic rhinosinusitis (CRS). (A) Reactome pathways enriched among CRSwNP-prioritized genes. (B) Gene Ontology (GO) biological processes enriched among CRSwNP-prioritized genes. (C) Reactome pathways enriched among CRS-prioritized genes. (D) GO biological processes enriched among CRS-prioritized genes. Each point represents a pathway, with the x-axis showing the enrichment odds ratio and the y-axis showing statistical significance (–log₁₀ p-value). Top pathways (highest –log₁₀ p-value) are labeled in each panel. Blue and red dots indicate pathways exceeding the significance threshold (e.g., FDR < 0.05); green dots fall below that threshold.

### Cell-type–specific expression patterns of GWAS-prioritized CRSwNP genes

Single-cell RNA-seq profiling of NP tissue detected 88 of the 92 GWAS-prioritized genes across 15 discrete cell-type clusters (**Table S12**). Differential expression analysis between epithelial and immune compartments (*FDR* < 0.05) identified 30 genes with significant cluster-specific enrichment. Seventeen genes—*GPX4, HINT1, ALOX15, RUNX1, LPP, NPEPPS, NDFIP1, ANK3, IL33, SMAD3, HDAC7, NELFE, TET2, STAT6, TMEM131, CYP2S1* and *GLIS3*—were preferentially expressed in epithelial lineages, whereas 13 genes—*RTF2, CD247, IL4R, RORA, RUNX3, FOXP1, ETS1, HLA-DQA1, IL7R, HLA-E, PTPRC, RPS26* and *RPS23*—were enriched in immune clusters (**Figure 3**). Furthermore, even within each broad compartment, expression varied by cell subtype: for example, *ALOX15* levels were highest in basal and goblet cells among epithelial subsets, and *IL7R* expression peaked in T cells relative to other immune subtypes (**Figure 3**).

**Figure 3.**
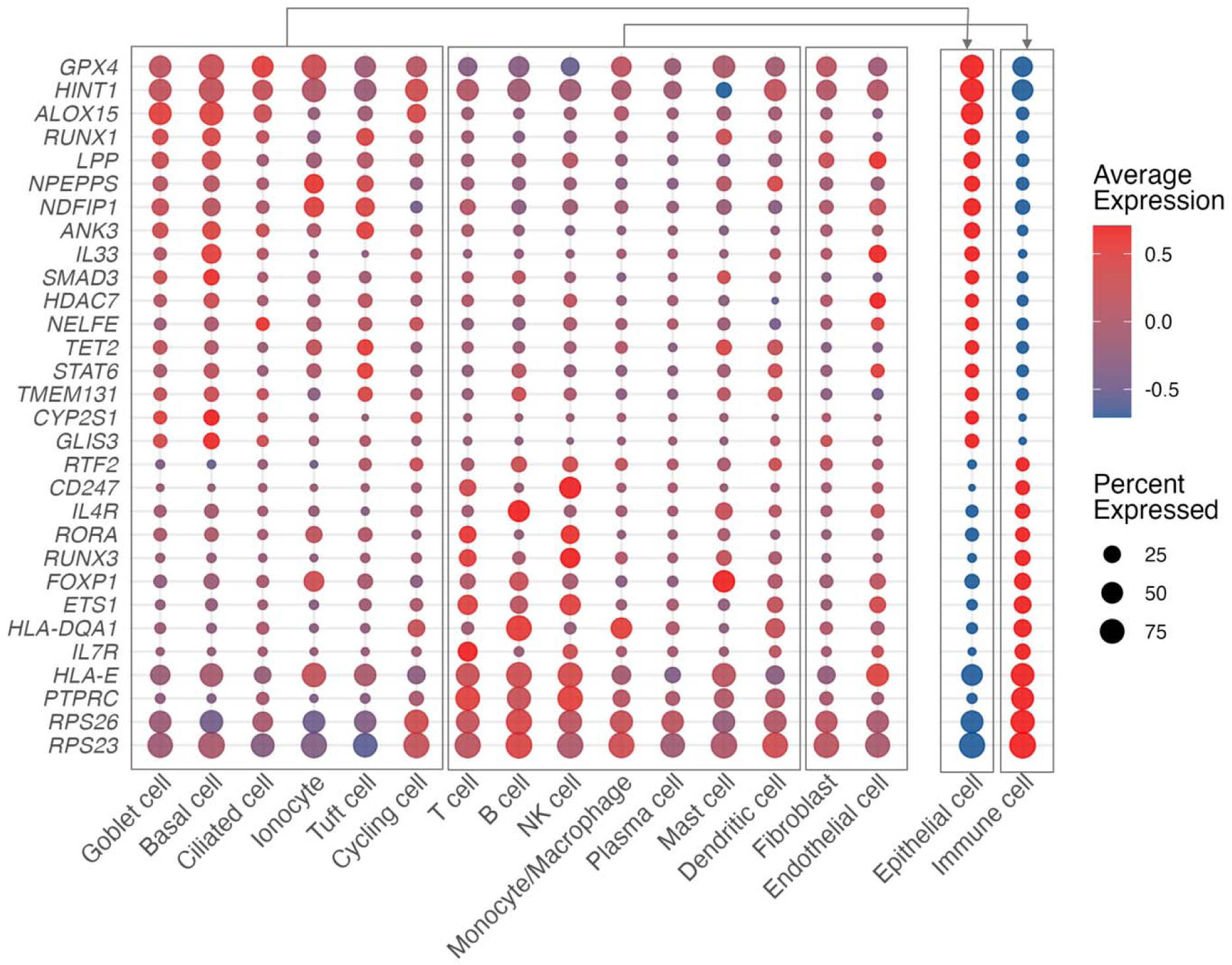
Single-cell transcriptomic profiling of chronic rhinosinusitis with nasal polyps (CRSwNP)-prioritized genes across epithelial and immune cell populations. Dot plot shows scaled average expression (color) and percentage of cells expressing each gene (dot size) for CRSwNP-associated genes with significant differential expression between epithelial and immune compartments. Cells are grouped by major nasal epithelial subsets (goblet, basal, ciliated, tuft, cycling, ionocyte) and immune subsets (monocytes/macrophages, plasma cells, dendritic, NK, T, B cells), with aggregate “Epithelial” and “Immune” columns at right. Dot color: scaled average expression (red = high, blue = low). Dot size: percent of cells expressing the gene (small = 25%, medium = 50%, large = 75%). Gene selection: only genes with FDRL<L0.05 for differential expression between epithelial versus immune cells are shown. Bracket: groups genes exhibiting epithelial-biased versus immune-biased expression patterns.

### Immune cells drive heritability enrichment of CRSwNP

We applied stratified LD score regression (LDSC-S) to partition CRSwNP heritability across 15 cell-type annotations, as well as aggregative epithelial and immune annotations (**Figure 4**). Among the 15 individual clusters, the strongest enrichments were observed in immune cell clusters—T cells (enrichment = 3.7; *P* = 0.002), NK cells (2.3; *P* = 0.002), B cells (2.8; *P* = 0.003), mast cells (2.6; *P* = 0.003), and dendritic cells (2.7; *P* = 0.015)—as well as within epithelial subsets, notably basal epithelial cells (2.0; *P* = 0.003), goblet cells (2.0; *P* = 0.036), and cycling cells (3.2; *P* = 0.048). When annotations were aggregated into broad immune versus epithelial categories, only the immune compartment remained significantly enriched (4.1; *P* = 0.010), whereas the epithelial compartment showed a non-significant trend (2.5; *P* = 0.072).

**Figure 4.**
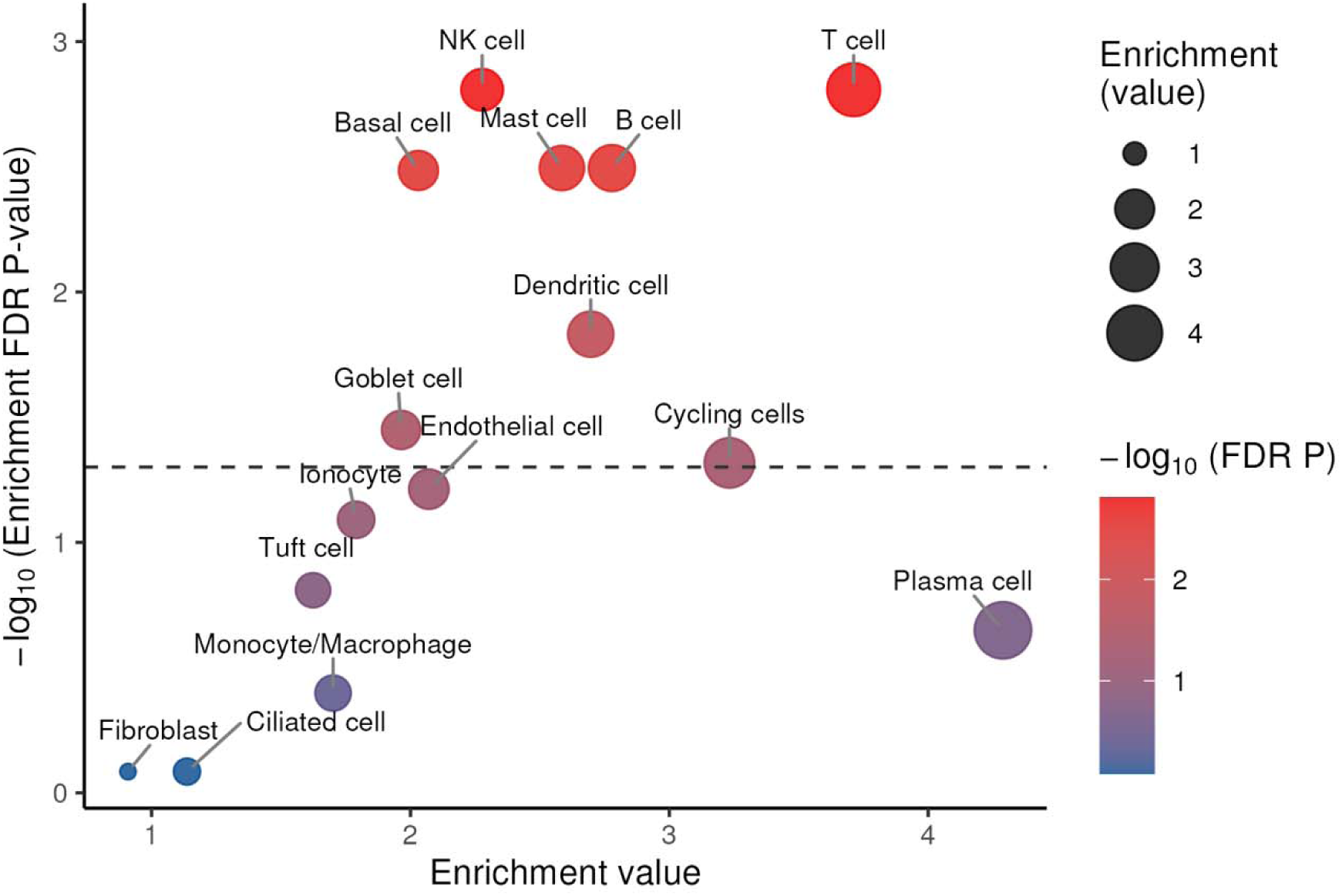
Partitional heritability of chronic rhinosinusitis with nasal polyps (CRSwNP) GWAS signals across single-cell populations. X-axis: fold-enrichment of CRSwNP GWAS SNP heritability within each annotation relative to the genome-wide baseline. Y-axis: statistical significance expressed as –log₁₀(FDR-adjusted P). Dot size scales with the enrichment value; dot color indicates –log₁₀(FDR P) (red = more significant). Horizontal dashed line marks FDR = 0.05; annotations above this line are considered significantly enriched. Cell-type labels highlight key epithelial (e.g., basal, goblet, tuft) and immune (e.g., T, NK, B, dendritic, plasma) populations with notable enrichment.

### Integrative genomic analyses reveal shared and distinct genetic architectures of CRSwNP and overall CRS

Significant local genetic correlations between CRSwNP and overall CRS were detected in 21 discrete genomic regions (FDR < 0.05), collectively encompassing 471 genes (**Figure 5a; Table S13**). CRSwNP-CRS colocalization then pinpointed 39 loci with high posterior probability of a shared causal variant (**Figure 5b; Table S14**). To further dissect shared versus disease-specific architecture, we implemented genomic SEM: a common-factor model capturing shared liability, alongside two GWAS-by-subtraction models isolating CRSwNP- and CRS-specific residual signals (**Figure 5c**). The common-factor SEM implicated 92 genome-wide loci mapping to 83 genes (**Table S15**), while the subtraction models identified three CRSwNP-specific and six CRS-specific loci (**Tables S16; Table S17**). Integrating all three methods (local rg, multi-trait colocalization, and genomic SEM) yielded 12 genes— *BACH2, CD247, FADS2, FOXP1, FUT2, GPX4, IL7R, NDFIP1, RAB5B, RORA, SMAD3, TSLP*—with consistent evidence for shared risk, plus 24 additional genes (e.g., *ALOX15, GATA3, RUNX3, IL2RB*) supported by two approaches (**Figure 5d; Table S18**).

**Figure 5.**
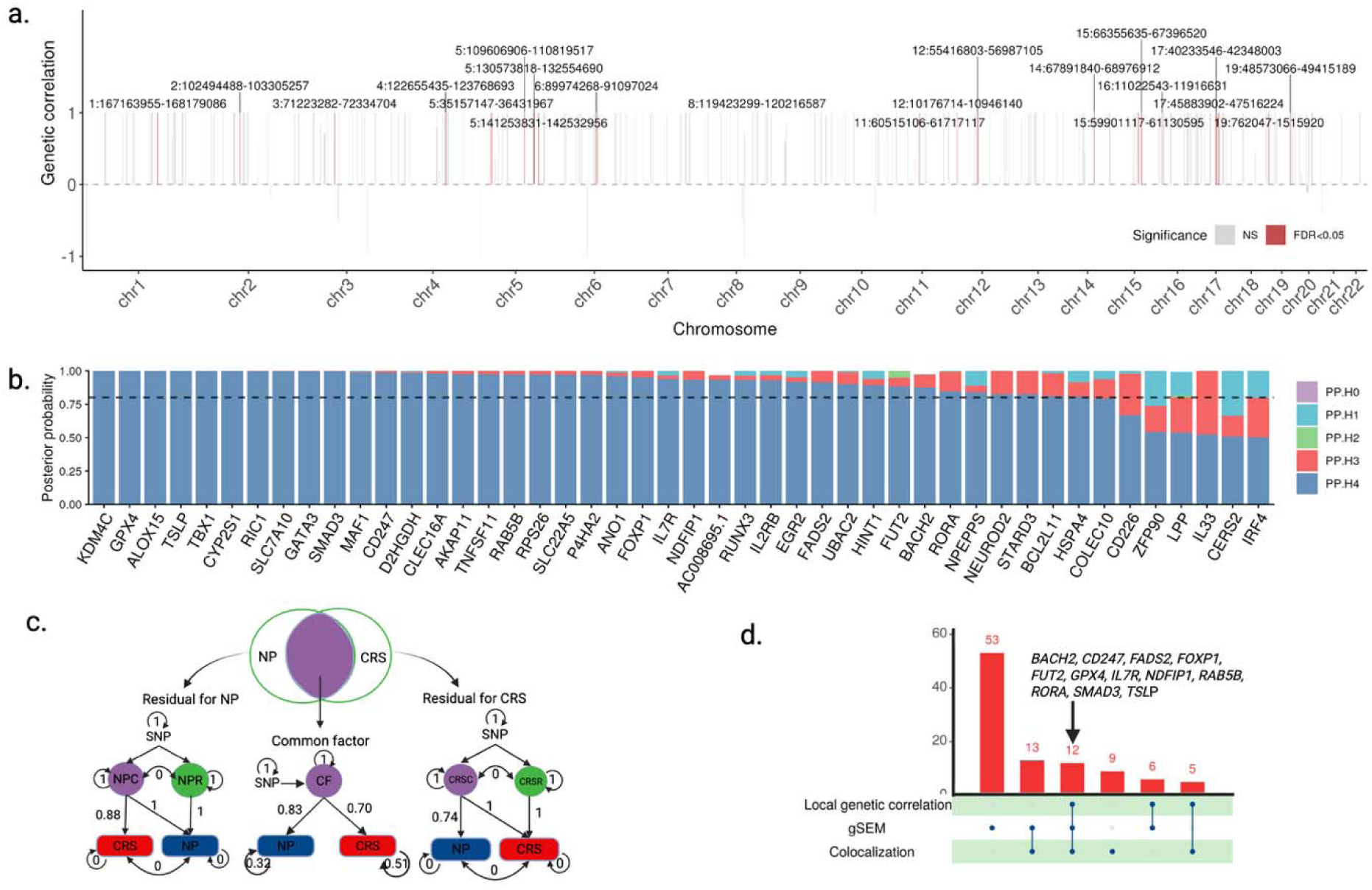
Shared genetic architecture of chronic rhinosinusitis with nasal polyps (CRSwNP) and overall chronic rhinosinusitis (CRS). (A) Local genetic correlation across the genome. Genome-wide map of local genetic correlation (ρ) between CRSwNP and CRS (hg38 coordinates). Bars above the axis indicate positive correlation; below, negative. Red bars denote regions with FDR < 0.05. Key significant loci are labeled by their chromosomal positions (chr:start–end). (B) CRSwNP-CRS colocalization posterior probabilities. (C) Genomic structural equation model (gSEM). Path diagram showing a common latent factor (purple) influencing both CRSwNP and CRS, plus trait-specific residual factors (green). Path coefficients (β) are indicated on arrows. (D) Overlap of shared genes identified by three methods. Bar plot showing the number of genes jointly implicated by (1) local genetic correlation (HDL), (2) colocalization, and (3) gSEM common factor GWAS loci. Genes detected by all three approaches are listed above the bars; the green row below marks which methods each gene passed.

### Protein-wide MR analysis reveals shared and unique blood proteins for CRSwNP and overall CRS

Across 2,864 proteins with available genetic instruments, SMR identified 80 whose genetically predicted levels were significantly associated with CRSwNP risk after FDR correction (**Figure 6a; Table S19**). Of these 80, 45 proteins could be tested in an independent pQTL GWAS: all but three showed directionally consistent effects, and 37 met nominal significance (*P* < 0.05) in the replication set (**Figure 6b; Table S20**). Bayesian colocalization with GWAS loci provided strong evidence (PPH4 > 0.70) for ten protein–CRSwNP pairs (**Figure 6c; Table S21**). In parallel, 34 proteins demonstrated FDR-corrected SMR associations with CRS (**Figure 6d; Table S22**), all of which were directionally concordant in the independent dataset and 30 reached nominal replication significance (**Figure 6e; Table S23**). Eight CRS–protein associations colocalized at PPH4 > 0.70 (**Figure 6f; Table S24**). When comparing across phenotypes, 23 proteins showed concordant SMR effects on both CRSwNP and CRS (**Table S25**), while three—TNFSF11, IL2RB and STX4—also exhibited strong colocalization support in both conditions.

**Figure 6.**
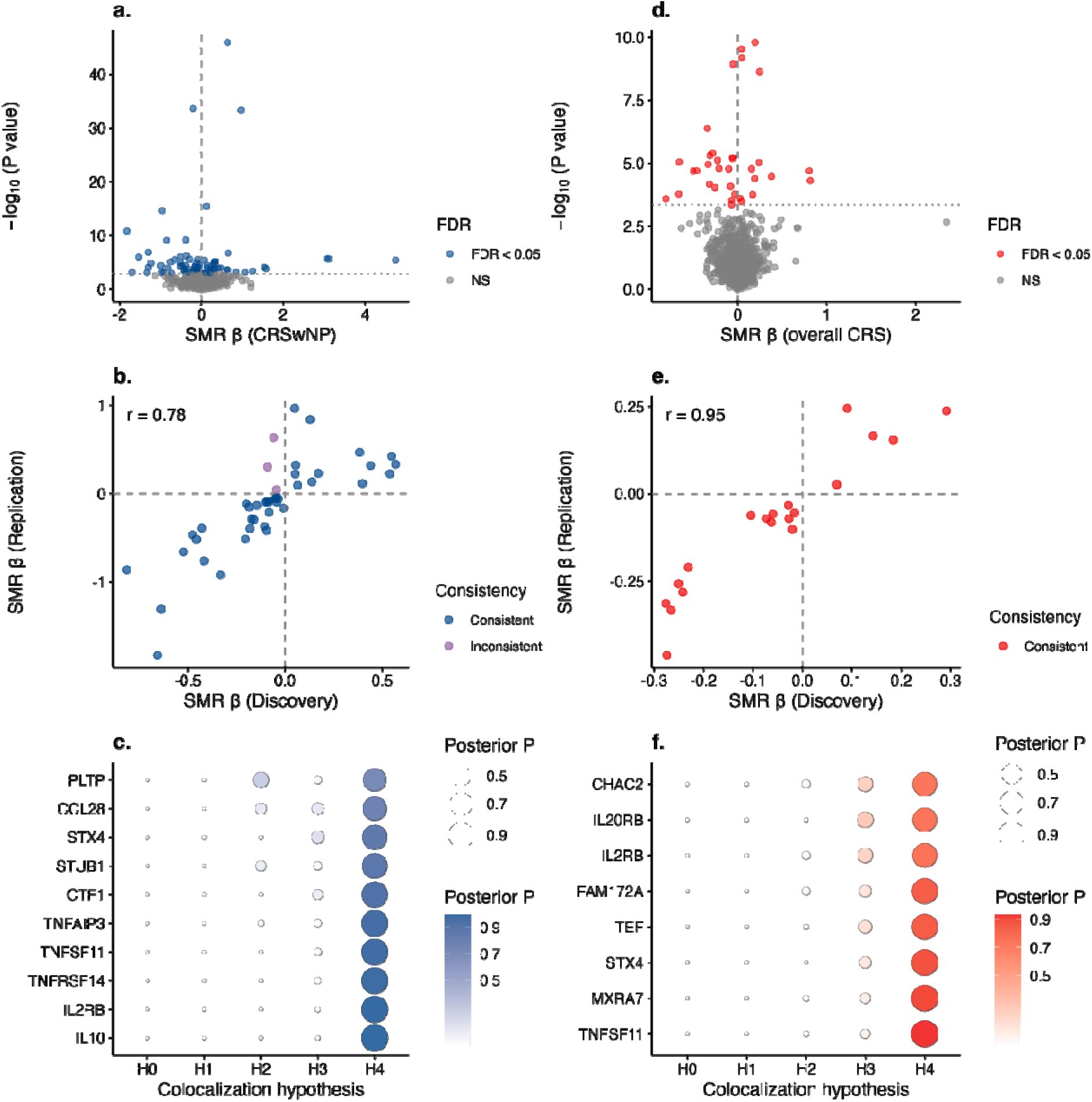
Mendelian randomization, replication, and colocalization of circulating proteins in relation to chronic rhinosinusitis with nasal polyps (CRSwNP) and overall chronic rhinosinusitis (CRS). (A) Two-sample protein-wide MR for CRSwNP risk. X-axis: causal estimate (log-OR per SD increase in protein level). Y-axis: statistical significance (–log₁₀ P for MR). Vertical dashed line: null effect; horizontal dashed line: FDRL=L0.05. (B) Replication of CRSwNP–protein associations in an independent pQTL GWAS. Dots colored in blue if replicated with concordant direction. (C) Colocalization between pQTLs and CRSwNP risk signals. Bubble size = posterior probability of a shared causal variant (PP.H4). Color intensity = PP.H4 (blue = highest). (D) Two-sample protein-wide MR for CRS risk, plotted as in (A). Axes and dashed lines as in (A). (E) Replication of CRS–protein associations in the same independent pQTL GWAS. (F) Colocalization between pQTLs and CRS risk signals, plotted as in (C). Color intensity = PP.H4 (red = highest).

## Discussion

This multi-ancestry GWAS meta-analysis mapped 96 genome-wide significant loci for CRSwNP and 41 for overall CRS, prioritizing 92 and 39 candidate genes, respectively. CRSwNP and overall CRS shared genetic susceptibility, but overall CRS exhibited broader genetic correlations across multiple respiratory disorders. Pathway enrichment consistently pointed to immune signaling albeit with disease-specific emphases and lipid-metabolism networks as key drivers of disease. At the single-cell level, CRSwNP-associated genes displayed distinct expression patterns across nasal epithelial and immune clusters, and immune annotations captured greater CRSwNP heritability than epithelial annotations. By integrating three comparative genetic analyses, we delineated multiple shared risk loci (e.g., *BACH2, CD247, FADS2, FOXP1, FUT2, GPX4, IL7R, NDFIP1, RAB5B, RORA, SMAD3, TSLP*) and a few disease-specific loci. Proteome-wide MR identified circulating proteins (e.g., TNFSF11, IL2RB and STX4) that influence both CRSwNP and overall CRS. Together, this comprehensive multi-omics framework prioritizes cell clusters in CRSwNP and defines shared versus disease-specific mechanisms in CRSwNP and overall CRS and nominates molecular targets for future therapeutic intervention. Earlier genetic studies, including family-based GWAS and modestly sized case-control analyses, implicated immune-related loci but lacked power for extensive locus discovery and detailed mechanistic inference (5,7). More recently, a UK Biobank and FinnGen study in individuals of European genetic ancestry identified 50 loci for CRSwNP and 5 loci for CRSsNP and reported substantial locus sharing between asthma and CRSwNP (8). Building on this work, our larger multi-ancestry meta-analysis identified 96 loci for CRSwNP and 41 loci for overall CRS, providing a higher-resolution view of genetic architecture and extending insights beyond the long-recognized contribution of the *HLA* region.

Several loci and prioritized genes, including *IL33, HLA, ALOX15*, *KDM4C*, *NELFE*, and *RIC1*, showed strong associations and point to more than one disease-relevant process, supporting biological heterogeneity within CRSwNP. These signals span epithelial cytokine pathways, antigen presentation, lipid mediator biology, and gene regulation or intracellular trafficking. *IL33* is consistent with an epithelial “alarmin” pathway that links barrier disruption to downstream type 2 inflammation, which has been highlighted as central to severe and treatment-refractory CRS/CRSwNP (9). The prominent associations at *HLA-DQA1* in CRSwNP and *HLA-C* in overall CRS further implicate antigen presentation and adaptive immune responses, in line with prior genetic and immunologic evidence for immune dysregulation in CRS/CRSwNP (5). *ALOX15* provides a direct connection to lipid mediator pathways (6), consistent with our enrichment of lipid metabolism processes shared across CRSwNP and CRS. Furthermore, ALOX15 is significantly elevated in patients with aspirin-exacerbated respiratory disease, a unique and particularly severe subtype of CRSwNP (10).In addition, candidates such as *KDM4C*, *NELFE*, and *RIC1* are plausible regulators of context-dependent gene expression and cellular state, which is particularly relevant for airway disease where exposures and the local tissue environment shape inflammatory programs (11). We observed lower SNP heritability and fewer loci for overall CRS than for CRSwNP. This pattern likely reflects the greater clinical and biological heterogeneity of overall CRS, which encompasses multiple endotypes and variable contributions from inflammation, infection, and structural factors, rather than indicating weaker biological underpinnings (9).

Consistent with the clinical and biological heterogeneity of overall CRS, we observed broader genetic correlations between CRS and respiratory disease outcomes than between CRSwNP and these outcomes. CRSwNP showed significant correlations with a smaller set of respiratory endpoints, with the strongest signal for asthma with exacerbation, whereas CRS was significantly correlated with a much larger fraction of endpoints, including bronchitis, asthma, and chronic airway obstruction. This pattern is clinically plausible: CRS likely reflects a spectrum of respiratory inflammation, which encompasses multiple endotypes and upstream risk pathways, with differential impact on the upper and lower respiratory systems. In contrast, CRSwNP appears to have a more focused genetic architecture, consistent with its higher SNP heritability and with nasal polyposis representing a more immune-centered condition within the broader CRS spectrum.

Pathway enrichment of the prioritized genes implicates immune signaling as a major component of risk for both CRSwNP and overall CRS, but with clear differences in the dominant programs. CRSwNP genes were enriched for cytokine-mediated pathways, particularly IL-4 and IL-13 signaling (12), whereas overall CRS genes showed stronger enrichment for T-cell–related processes and IL-1 signaling (13). This pattern is consistent with current endotype frameworks in which nasal polyposis is frequently associated with type 2 inflammation, while CRS includes a wider range of inflammatory mechanisms. In addition, both CRSwNP and CRS showed enrichment of lipid metabolism pathways involving eicosapentaenoic and arachidonic acid processes. These signals are most consistent with lipid mediator and immunometabolic pathways that influence inflammation (14,15), rather than with lipid transport pathways typical of cardiometabolic disease (16). The strong CRSwNP association at *ALOX15* supports this interpretation by linking locus-level evidence to the shared enrichment of lipid mediator biology (6).

Integration of CRSwNP GWAS signals with single-cell RNA-seq found that several genes were preferentially expressed in epithelial subsets, including *IL33* and *ALOX15*, whereas others were enriched in immune clusters, including *CD247*, *IL4R*, *FOXP1*, *HLA-DQA1*, and *IL7R*. These patterns support a two-compartment model in which epithelial programs can initiate or amplify inflammatory signals, while immune programs help maintain chronic inflammation. Heritability partitioning provided complementary evidence that inherited CRSwNP risk is concentrated in immune cell annotations. Multiple immune clusters, including T cells, NK cells, B cells, mast cells, and dendritic cells, showed significant enrichment, and only the aggregated immune compartment remained significant. This immune-enriched architecture is consistent with the clinical effectiveness of immune-directed therapies in severe nasal polyposis (17) and suggests that germline susceptibility acts mainly through immune regulation rather than through purely structural epithelial mechanisms.

The presence of some epithelial signals is not inconsistent with immune-skewed heritability enrichment. Instead, our findings support a conservative and testable model in which germline variation sets immune activation thresholds, whereas epithelial remodeling and barrier dysfunction are more dependent on disease state (18) and exposures (19), such as allergens, microbial factors, pollutants, and smoking. These exposures may act through persistent inflammatory signaling and epigenetic changes that stabilize disease-relevant epithelial programs (20). We emphasize that this is a hypothesis generated by compartment-specific expression and enrichment patterns; direct evidence will require longitudinal tissue-based multi-omics and context-specific QTL analyses in nasal epithelium.

Our comparative analyses identified shared genetic signals between CRSwNP and overall CRS that point to a biologically coherent core liability that likely acts upstream of current diagnostic labels. A central theme is epithelial licensing together with adaptive immune control. On the epithelial side, *TSLP* and *IL1RL1* implicate alarmin signaling as a shared gateway that can amplify downstream inflammation in the setting of persistent barrier stress and antigen exposure. Importantly, this axis is supported by clinical trial evidence: tezepelumab (anti-TSLP) improved nasal polyp burden and sinonasal symptoms and reduced the need for surgery or systemic glucocorticoids in the phase 3 WAYPOINT trial (21), and it is now approved by the U.S. FDA as add-on maintenance treatment for inadequately controlled CRSwNP.

On the immune side, the shared module is anchored by *CD247* (T-cell receptor signaling) and *IL7R* (T-cell survival and homeostasis), together with transcriptional regulators that shape lymphocyte fate and polarization (*FOXP1, BACH2, RUNX3, RORA, GATA3*). This architecture is consistent with biobank-scale studies showing shared genetic components between asthma and CRSwNP and enrichment of immune pathways across these conditions (8). The therapeutic relevance is further supported by the fact that downstream nodes on the same type 2 immune axis already improve clinical outcomes in randomized trials in CRSwNP, including dupilumab (IL-4R alpha) (22), mepolizumab (IL-5) (23), and omalizumab (IgE) (24). Together, these lines of evidence support the inference that shared loci are enriched for pathways with demonstrable interventional leverage, consistent with a causal immune and epithelial biology rather than secondary tissue remodeling alone.

Beyond cytokine signaling, the shared genes also highlight mechanistic layers that may contribute to heterogeneity and variable treatment response. First, an immunometabolic and lipid-redox component, including *FADS2, ALOX15, GPX4,* and *CYP2S1*, is consistent with enrichment of arachidonic and eicosapentaenoic acid pathways and suggests that variation in lipid mediator generation and oxidative resilience may modify mucosal inflammation in CRSwNP and overall CRS (14,15). Second, shared candidates involved in intracellular trafficking and ubiquitin-linked regulation (*RAB5B, NDFIP1, RIC1, UBAC2, CLEC16A*) plausibly influence receptor recycling, antigen handling, and secretory dynamics (25). These process-level mechanisms could contribute to persistence and recurrence and may not be captured by bulk cytokine measures (26), providing a rationale for future work on endotype stratification and combination strategies.

MR analysis showed that genetically predicted higher circulating levels of TNFSF11, IL2RB, and STX4 were associated with lower risk of both CRSwNP and overall CRS. This consistent protective direction helps prioritize these proteins as candidate mediators and also informs the expected direction of therapeutic perturbation. For TNFSF11 (RANKL), the finding is notable because RANKL inhibition with denosumab is widely used in clinical practice. Although our genetic results do not establish drug causality for CRS, the U.S. prescribing information for Prolia notes that RANKL is expressed on activated T and B lymphocytes and that RANKL inhibition may increase infection risk (27), with upper respiratory tract infections reported among adverse events. These label statements support a plausible pharmacovigilance hypothesis that sustained RANKL blockade could, in some settings, shift susceptibility to upper airway infection or inflammation in a direction that is consistent with TNFSF11 being protective in genetic analyses. For IL2RB, the protective association aligns with the broader theme of immune set-point regulation but has complex translational implications because IL-2/IL-15 signaling can have different effects across immune cell subsets (28). STX4 is less directly druggable, but its protective association supports a role for trafficking and secretion processes in disease biology, consistent with other shared loci that implicate intracellular transport and signaling control.

The genomic SEM GWAS-by-subtraction models identified a small number of loci with residual associations that were not well explained by the common CRSwNP–CRS factor, including 3 CRSwNP-specific and 5 CRS-specific loci. For CRSwNP, two loci mapped near UXS1 and SIX4, nominating processes that may be more closely linked to tissue remodeling and local epithelial biology (29). For overall CRS, the subtraction signals included loci near TNFRSF14, TMEFF2, UGT2A2, and PPP1R12A, spanning immune signaling, epithelial state regulation, mucosal metabolism, and cytoskeletal control. In particular, UGT2A2 is highly expressed in nasal mucosa and has established roles in local xenobiotic and endogenous metabolite handling (30), supporting a plausible CRS-leaning mechanism that reflects sinonasal epithelial physiology. However, these subtraction-model loci should be interpreted cautiously. Apparent disease specificity can be inflated by differences in phenotype definitions and ascertainment across cohorts (31). In addition, GWAS-by-subtraction relies on assumptions about the shared latent factor and can have reduced power or increased sensitivity to misclassification when traits are strongly correlated (32), so residual signals may reflect a mixture of true disease-specific biology and modeling or phenotyping artifacts.

Several limitations should be considered when interpreting these results. First, phenotype definitions differed across cohorts and health systems, which likely reduced power by introducing heterogeneity and may have obscured endotype-specific signals, particularly for overall CRS. Second, although we used a multi-population design, sample sizes were not balanced across ancestry groups, which limits inference for ancestry-specific effects and reduces confidence in generalizability for underrepresented populations. Third, gene prioritization and pathway enrichment are inherently indirect and probabilistic; assigning causal genes at individual loci will require targeted functional follow-up. Fourth, single-cell expression mapping and heritability partitioning help prioritize relevant cellular compartments but do not establish causal cell types and may be influenced by disease stage, sampling context, and treatment exposure. Finally, MR analyses can be affected by horizontal pleiotropy and by differences between circulating protein levels and local tissue activity, even when colocalization supports a shared genetic signal.

This multi-population GWAS meta-analysis expanded genetic discovery for CRSwNP and overall CRS and showed substantial shared liability with distinct disease-specific components. Integrative analyses prioritize a shared epithelial alarmin–immune regulatory module. These findings provide a mechanistic framework for endotype-based stratification and therapeutic target prioritization in chronic sinonasal inflammation.

## Methods

We used NP codes as a proxy for CRSwNP and overall CRS codes as a broader, heterogeneous phenotype likely capturing both CRSwNP and CRSsNP in unknown proportions (33,34). On this basis, we carried out a trans-population GWAS meta-analysis of CRSwNP and overall CRS by harmonizing International Classification of Disease (ICD)- and phecode-based case definitions across six large biobanks (FinnGen (35), UK Biobank (36), MVP (37), All of Us (38), Genes & Health (39) and BioBank Japan (40)), spanning European, African, Admixed American, South Asian and East Asian populations. In total, 17 046 individuals with CRSwNP and 1,573,214 controls without CRSwNP, as well as 97,519 individuals with overall CRS and 1 612 702 controls without CRS, were included. All research participants provided informed consent, and ethical approval was obtained from the relevant institutional review boards. Details of the included cohorts, including recruitment strategies, case–control definitions, genotyping, imputation, and GWAS procedures, are provided in the **Supplementary Methods**. Datasets reported in hg19 were converted to hg38 using liftover tools (41). GWAS summary statistics were then assessed for quality using GWASinspector (42) prior meta-analysis. We evaluated trans-ancestry genetic architecture using the Popcorn Python package (43) to quantify cross-population effect heterogeneity and no significant ancestry-specific differences were detected. Variants with a minor allele count (MAC) <50 were excluded from individual summary statistics. Meta-analysis was conducted using METAL (44) in standard error mode with genomic control correction applied to each cohort. Following meta-analysis, variants present in only one cohort or with minor allele frequency (MAF) < 1% were excluded from downstream analyses. We applied Linkage Disequilibrium Score Regression (LDSC) (45) to quantify the extent of polygenic signal versus confounding in our GWAS and to estimate SNP-based heritability on the liability scale for CRSwNP (prevalence = 4%) and overall CRS (prevalence = 12%)

### Genetic correlations

Genome-wide genetic correlations (*r_g_*) were estimated using LDSC (45). GWAS summary statistics for each phenotype were limited to HapMap3 SNPs (MAF > 1%, INFO > 0.9), and LD scores were computed from the 1000 Genomes Project Phase 3 European reference panel. We first conducted bivariate LDSC with unconstrained intercepts to account for sample overlap, deriving the genetic correlation between CRSwNP and overall CRS. To contextualize these findings, the same pipeline was applied to estimate pairwise *r_g_* values between CRSwNP or overall CRS and a spectrum of respiratory-related traits in the MVP cohort (37) (e.g., asthma, chronic obstructive pulmonary disease, pulmonary fibrosis). Bonferroni correction was applied across all tested trait pairs to control for multiple comparisons.

### Definition of loci

Genome-wide significant SNPs (*P* < 5 × 10⁻□) were grouped into loci based on physical proximity. Loci were defined by: (1) identifying all genome-wide significant variants from the association results; (2) extending 250 kb upstream and downstream of each variant to create 0.5 Mb windows; and (3) merging overlapping windows to form distinct loci (46). For each locus, the SNP with the lowest *P* value was designated as the index variant.

### Gene prioritization

We applied five complementary gene prioritization approaches: (1) nearest gene annotation, (2) Polygenic Priority Score (PoPS) (47), (3) expression quantitative trait loci (eQTL) colocalization, (4) functional annotation based on CARMA (Credible-variant Analysis for Regional Meta-Analysis) (48), and (5) chromatin accessibility annotation. For each genomic locus, genes were prioritized based on the total number of times they were selected across these methods. In cases where multiple genes had the same selection count, we refined prioritization by first considering genes containing variants within CARMA-identified credible sets with functional annotation; if ambiguity remained, the nearest gene to the index variant was selected.

1. **Nearest gene annotation** For each locus, the lead SNP was assigned to the gene whose body lay at the shortest physical distance from the variant. Nearest-gene mapping was carried out with the get_nearest_gene() function from the gwasRtools R package (https://github.com/lcpilling/gwasRtools).
2. **PoPS** We first applied MAGMA (Multi-marker Analysis of GenoMic Annotation) (49) to derive gene-level association statistics by aggregating SNP-level summary data within each gene’s boundaries and adjusting for local linkage disequilibrium. We then used PoPS, a similarity-based prioritization framework that integrates these MAGMA metrics with diverse functional annotations—such as RNA-seq expression profiles, curated pathway databases, and predicted protein–protein interaction networks—to select the most informative features and compute a causality score for each gene. Under the assumption that causal genes share common functional signatures, all genes within ±1 Mb of each lead variant were ranked by their PoPS scores, and the highest-scoring gene at each genome-wide significant locus was designated as the top candidate (47).
3. **eQTL colocalization** Colocalization analyses were performed using the coloc R package’s coloc.abf() function (https://github.com/chr1swallace/coloc), which applies an approximate Bayes factor framework to determine whether two traits share a causal variant (50). We specified priors of p₁ = 1×10⁻□, p₂ = 1×10⁻□and p₁₂ = 1×10⁻□to calculate posterior probabilities for the five standard hypotheses—no association with either trait (H₀), association with only one trait (H₁ or H₂), association with both traits but different causal variants (H₃), and association with both traits via a shared causal variant (H₄)—and considered PP₄ > 0.7 as evidence of colocalization. For each GWAS index SNP, all variants within ±500 kb were extracted and tested using eQTLGen Phase I data (51).
4. **Functional annotation of fine-mapped variants** We employed CARMA (48), a Bayesian fine-mapping framework that models local linkage disequilibrium and integrates summary-level association signals across studies or populations to estimate each variant’s posterior inclusion probability. CARMA ranks variants by their posterior probabilities and constructs minimal credible sets—here defined as the smallest set of variants whose cumulative posterior probability reaches 95%—thereby pinpointing those most likely to be causal at each locus. All variants within these 95% credible sets were functionally annotated via the Open Targets platform (https://www.opentargets.org/), which provides coding, regulatory, and splicing effect predictions (52). When a variant was predicted to directly impact a gene’s function—either by altering its coding sequence or regulatory elements—that gene was designated as the candidate effector at the corresponding locus.
5. **Open chromatin accessibility annotation** To annotate regulatory potential, we first extracted SNPs from the 95% credible sets identified by CARMA fine mapping. These variants were then overlapped with open chromatin regions profiled in primary mononuclear cells from peripheral blood (E062) in the NIH Roadmap Epigenomics Project (53), using ChIP-seq peaks for H3K27ac (active enhancers and promoters), H3K4me1 (poised and active enhancers), and H3K4me3 (active promoters). Variants residing within these marked regions were subsequently linked to candidate target genes via the Activity-by-Contact (ABC) model, which integrates chromatin accessibility with three-dimensional contact frequency to predict enhancer–gene interactions (54). This strategy enabled the assignment of credible SNPs to their most likely effector genes based on tissue-specific regulatory architecture.

### Pathway enrichment

Pathway enrichment analyses were conducted to identify biological pathways and functional categories significantly overrepresented among the prioritized genes. Enrichment was performed using the enrichR R package (55) against the Reactome and Gene Ontology biological processes databases. Multiple testing was controlled by the Benjamini–Hochberg false discovery rate, with adjusted *P* < 0.05 deemed significant.

### Cell-type expression of candidate genes

Single cell RNA sequencing (scRNA-seq) data were extracted from nasal tissues from 9 patients with NP with data publicly available (**Figure S1a**) (56–58). Raw count matrices, gene annotations, and barcode information were downloaded from the NIH Gene Expression Omnibus (GEO; https://www.ncbi.nlm.nih.gov/geo/) under accession numbers GSE202100, GSE156285, and GSE179269. Data processing and analysis were conducted using the Seurat R package (version 5.0.2; https://satijalab.org/seurat/) (59) in R version 4.3.3. Cells were retained if they expressed between 250 and 2000 genes. Cells with fewer than 200 or more than 5000 total reads were excluded, as were cells in which more than 20% of reads mapped to mitochondrial genes. Mitochondrial content was calculated using the PercentageFeatureSet function with the pattern “^MT-”. Following quality control, 28,780 cells remained for downstream analysis. Data were normalized using the NormalizeData function with the LogNormalize method and a scaling factor of 10^4^. The top 2,000 highly variable genes were identified using the “vst” selection method. Genes were then scaled and centered (mean = 0) using the ScaleData function. Principal component analysis was performed using the RunPCA function based on these variable genes. To correct for batch effects, the Harmony package (version 1.2.0) (60) was applied. Cell clustering was performed using the FindNeighbors and FindClusters functions with a resolution parameter set to 0.5. To visualize the clusters in two dimensions, nonlinear dimensionality reduction was applied using the RunUMAP function. Cluster**-**specific marker genes were identified using the FindAllMarkers function by comparing gene expression in each cluster against all remaining clusters. Manual inspection of these differentially expressed genes (DEGs) was used to assign cluster identities (**Figure S1b** and **Figure S1c**). DEG analysis between selected groups as well as between epithelial (goblet cells, basal cells, ciliated cells, ionocytes, cycling cells and tuft cells) and immune (T cell, monocyte/macrophage, B cell, mast cell, plasma cell, NK cell, and dendritic cell) cell types were conducted using the FindMarkers function. Genes were considered differentially expressed if they exhibited an adjusted *P* value < 0.05 and a log2 fold change > 0.8 or < –0.8. The average expression and percent expressed estimates were calculated for NP genes against 15 single cell clusters as well as epithelial and immune cell clusters. Seurat’s DotPlot() was used for visualization.

### Cell-type inheritability

To quantify cell-type-specific heritability enrichment, we performed stratified LD score regression (LDSC-S) (61) using annotations derived from our scRNA-seq data. First, differentially expressed genes were identified for each annotated cell cluster via Wilcoxon rank-sum tests on log-normalized counts (*P* < 0.05). For each cell cluster, we then constructed a binary annotation file by extracting the genomic coordinates of each differentially expressed genes (gene body only) and selecting 1000 Genomes Phase 3 SNPs that fall within those boundaries. Cell-type–specific LD scores were generated with the ldsc.py tool by integrating these annotation tracks alongside the standard baseline LD model. Finally, we applied partitioned heritability analysis to our CRSwNP GWAS summary statistics, yielding enrichment coefficients that pinpointed which cell-type–defined gene sets contribute disproportionately to trait heritability.

### Common genetic components between CRSwNP and CRS

1. **Local genetic correlations** Local genetic correlations were estimated using the HDL-L framework, an extension of high-definition likelihood for locus-specific analysis (62). Briefly, the genome was partitioned into approximately independent LD blocks, and GWAS summary statistics for CRSwNP and overall CRS were harmonized to the 1000 Genomes Phase 3 European reference panel. Within each block, HDL-L models the local LD structure to derive maximum-likelihood estimates of genetic variance for each trait (h_1_ ^2^ and h_2_ ^2^) and their genetic covariance (Cov_g_). The local genetic correlation (*r_g, local_*) for each block was then calculated as

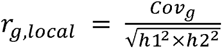

Block-specific standard errors for *r_g, local_* were obtained from the HDL-L likelihood profiles, and significance was assessed after Bonferroni correction across all tested blocks. To facilitate mechanistic inference, we extracted all genes whose genomic coordinates overlapped the identified LD blocks using GenomicRanges R package (63).
2. **CRSwNP-CRS colocalization analysis** We applied colocalization analyses coloc.abf() function to identify shared causal variants between CRSwNP and overall CRS. For each locus, we extracted GWAS summary statistics for both traits across all variants in the region. Using the same analytical setting as previously described, we then ran colocalization and obtained posterior probabilities for hypothesis 0 to 4 (PP.H0 to PP.H4). Loci with PP.H4 ≥ 0.8 were considered to harbor a single variant driving both CRSwNP and overall CRS associations, thus pinpointing shared signals.
3. **Genomic structural equation modelling (SEM)** We implemented Genomic SEM using the GenomicSEM R package (32) to decompose shared versus trait-specific genetic architectures for CRSwNP and overall CRS. First, we fitted a common factor model in which a single latent factor captures the genetic covariance between CRSwNP and overall CRS, identifying loci whose effects load significantly on this shared factor. Next, we specified two subtraction models—one for CRSwNP and one for overall CRS—by regressing each phenotype on the common factor and estimating residual genetic effects. Genome-wide significant loci emerging in the CRSwNP (or CRS) subtraction model but not reaching significance in the common factor model were classified as CRSwNP-specific (or CRS-specific) loci. Through this three-model framework, we distinguished pleiotropic variants influencing both diseases from those uniquely associated with each condition.

### Protein-wide Mendelian randomization

Protein-wide Mendelian randomization (MR) (64) association analyses were conducted against CRSwNP and overall CRS using European data. Genetic instruments for circulating proteins were selected from two sources: the UK Biobank Pharma Proteomics Project (UKB-PPP) (65) and deCODE (66). UKB-PPP analyzed 2,940 unique circulating proteins in 54,219 individuals using the Olink platform, while deCODE examined over 4,970 plasma proteins in 35,559 Icelanders using the SomaScan platform. For proteins with identical UniProt IDs in deCODE, one protein was retained based on stronger associations with the outcome. Overlapping proteins between the two datasets were sourced from UKB-PPP due to its larger sample size. The final analysis included 2,864 proteins with available genetic instruments that were the lead genetic variants associated with the proteins at the genome-wide significant level (**Figure S2**). The Fenland study (67), which measured 3,892 proteins in 10,708 individuals using the SomaScan platform, served as an independent replication dataset (**Figure S2**). Summary-based Mendelian randomization (SMR) (68) was applied to assess the associations of genetically predicted protein levels with CRSwNP and overall CRS risk, estimating odds ratios (ORs) per standard deviation (SD) increase in protein concentration. To account for multiple comparisons, *P*-values were adjusted using the false discovery rate (FDR), and associations with FDR-corrected *P*lJ<lJ0.05 were considered significant. Complementary Bayesian colocalization analysis was then performed to evaluate whether the same causal variant underlies the protein QTL and the disease association; posterior probability for hypothesis 4 (PPH4)lJ>lJ0.70 was taken as strong evidence of a shared causal signal.

## Data availability

The GWAS data generated in this study will be deposited in the NHGRI-EBI GWAS catalog database once the paper is published online.

## Acknowledgments

S.Y. was supported by an Award from the American Heart Association and the VIVA Foundation (https://doi.org/10.58275/AHA.24POST1189614.pc.gr.190880; Award ID: 24POST1189614). J.C.M. was supported by the National Institutes of Health grants T32-CA251063.

## Authors’ contributions

S.Y., M.G.L., and S.M.D. conceived and designed the study. S.Y. and J.C.M. contributed to data collection. S.Y. and J.C.M undertook the statistical analyses. S.Y. created the data visualizations and authored the initial draft of the manuscript. S.Y., J.C.M., XXX, B.F.V., M.G.L., and S.M.D. contributed to data interpretation, offered significant intellectual insights to the manuscript, and approved its final version.

## Statement of conflicts

S.M.D. receives personal consulting fees from Tourmaline Bio, research support from RenalytixAI, and in-kind research support from Novo Nordisk and Amgen, all outside the scope of the current project. Other authors declare no conflicts of interests.

## Notes

### Funding Statement

This study did not receive any funding.

### Author Declarations

This study only used publically available data sources. All data used in the study have been cited.

