## Supplementary Figures for "Comparative genomic analysis reveals shared and distinct mechanisms of nasal polyps and chronic rhinosinusitis"

Supporting information for

**Single-cell characterization of nasal polyps and comparative genomic analysis revealing shared and unique mechanisms with chronic rhinosinusitis**

Shuai Yuan, Jack C McVey, Katherine Hartmann, Sarah Abramowitz, Jakob Woerner, XXXX, Benjamin F. Voight, Michael G. Levin, Scott M. Damrauer

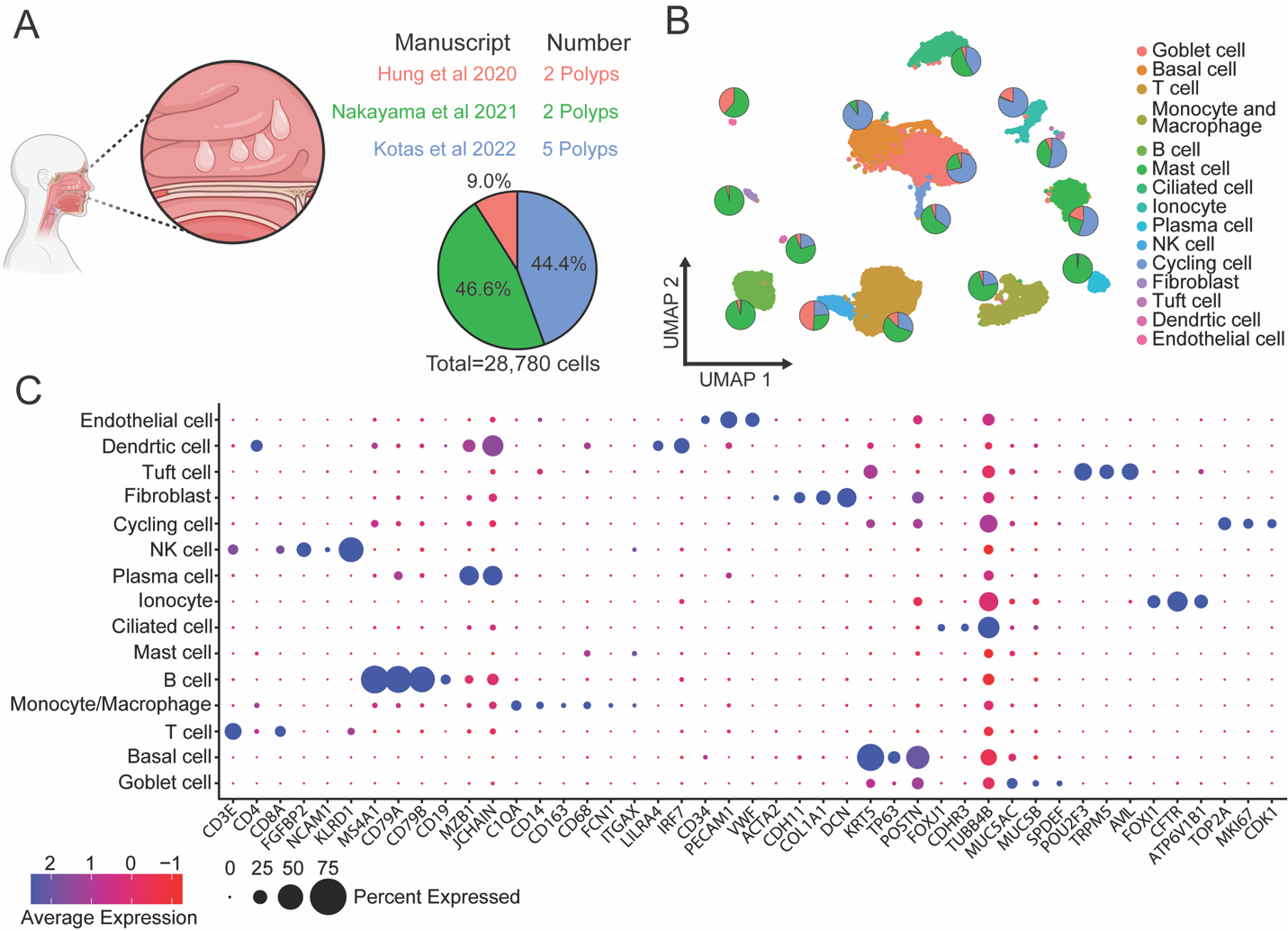

**Figure S1. Single cell analyses among nasal polyps**. Panel a. sample information. Panel b. Umap of cell types derived from 9 nasal polyps’ samples. Panel c. key gene expression across cells.

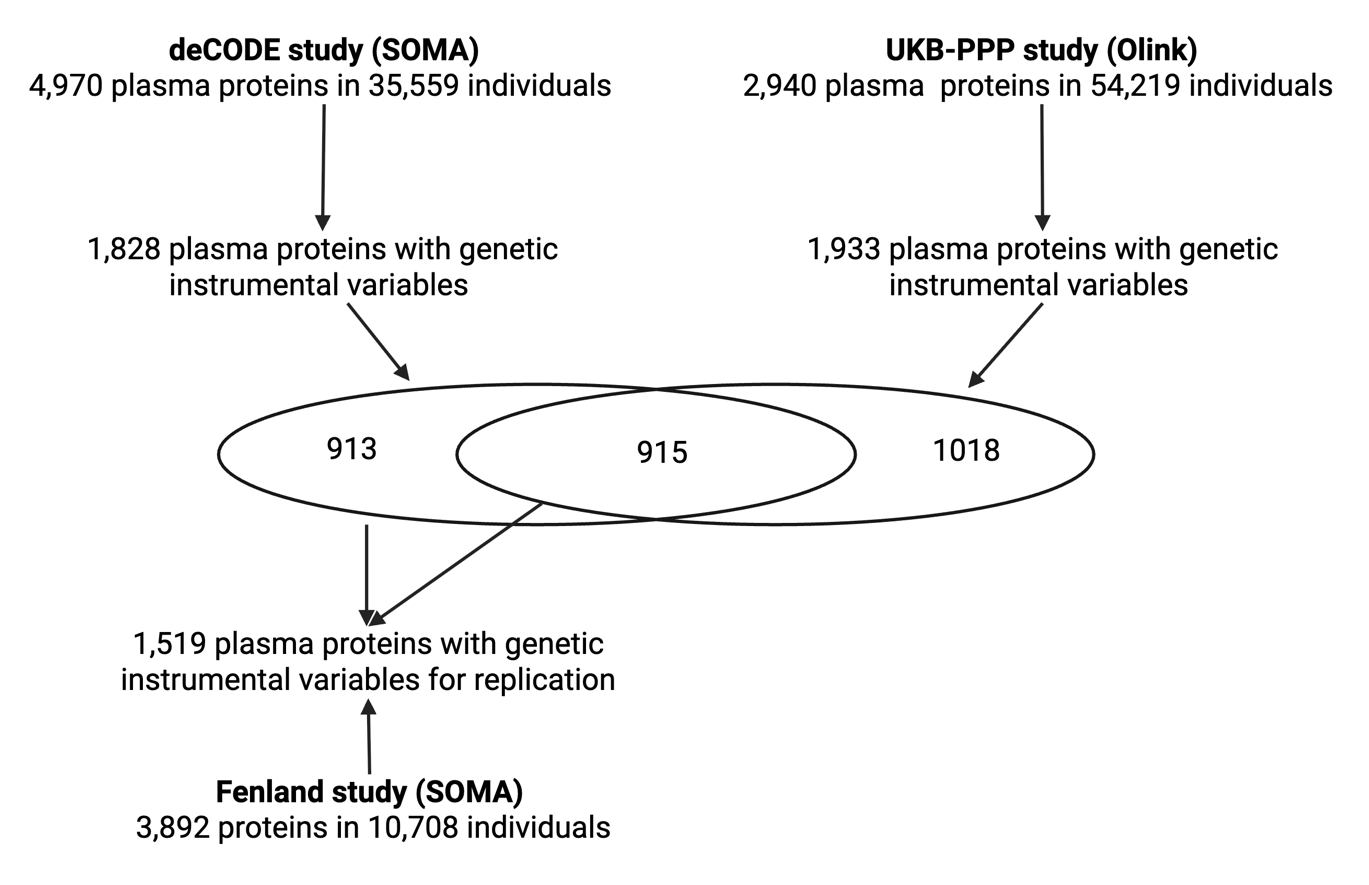

**Figure S2. Blood protein pool for Mendelian randomization analysis**. The primary analysis used data from deCODE and UKB-PPP studies after removal of duplicated proteins. The Fenland study was used as the replication study.

### **Supplementary tables 1-25 (see supplement excel results)**

#### Table S1. Study included in GWAS meta-analysis

#### Table S2. Genetic loci associated with nasal polyps at the genome-wide significance level in GWAS meta-analysis

#### Table S3. Genetic loci associated with chronic rhinosinusitis at the genome-wide significance level in GWAS meta-analysis

#### Table S4. Gene prioritization for nasal polyps genetic loci

#### Table S5. Gene prioritization for chronic rhinosinusitis genetic loci

#### Table S6. Genetic association between nasal polyps and other respiratory diseases in the VA MVP

#### Table S7. Genetic association between chronic rhinosinusitis and other respiratory diseases in the VA MVP

#### Table S8. Reactome pathway enrichment based on nasal polyps loci

#### Table S9. Gene Ontology (GO) biological process enrichment based on nasal polyps loci

#### Table S10. Reactome pathway enrichment based on chronic rhinosinusitis loci

#### Table S11. Gene Ontology (GO) biological process enrichment based on chronic rhinosinusitis loci

#### Table S12. Gene expression across single cells among nasal polyps samples

#### Table S13. Significant local genetic correlations between nasal polyps and chronic rhinosinusitis

#### Table S14. Genetic colocalization between nasal polyps and chronic rhinosinusitis

#### Table S15. Genetic loci associated with common factor generated by genomic structural equation modelling

#### Table S16. Genetic loci associated with nasal polyps residual generated by genomic structural equation modelling

#### Table S17. Genetic loci associated with chronic rhinosinusitis residual generated by genomic structural equation modelling

#### Table S18. Shared genes between nasal polyps and chronic rhinosinusitis in three comparative analyses

#### Table S19. Mendelian randomization association between blood proteins and risk of nasal polyps in discovery analysis

#### Table S20. Mendelian randomization association between blood proteins and risk of nasal polyps in replication analysis

#### Table S21. Genetic colocalization between blood protein levels and nasal polyps

#### Table S22. Mendelian randomization association between blood proteins and risk of chronic rhinosinusitis in discovery analysis

#### Table S23. Mendelian randomization association between blood proteins and risk of chronic rhinosinusitis in replication analysis

#### Table S24. Genetic colocalization between blood protein levels and chronic rhinosinusitis

#### Table S25. Consistent Mendelian randomization association between blood protein and nasal polyps and between blood protein and chronic
